# Corticolimbic network perturbations in clinical high-risk and first-episode psychosis: an arterial spin labelling and [^11^C]Ro15-4513 positron emission tomography study

**DOI:** 10.1101/2025.09.11.25335038

**Authors:** Samuel R. Knight, Julia J. Schubert, Mario Severino, Paulina B. Lukow, Amanda Kiemes, Nicholas R. Livingston, Fernando Zelaya, Andrea De Micheli, Zerrin Atakan, James Davies, Thomas J Spencer, Paolo Fusar-Poli, Jacek Donocik, Natasha Vorontsova, Eugenii A. Rabiner, Anthony A. Grace, Federico E. Turkheimer, Steven C.R. Williams, Mattia Veronese, Owen O’Daly, Philip McGuire, Gemma Modinos

## Abstract

Hippocampal regional cerebral blood flow (rCBF) alterations in early psychosis may result from GABAergic inhibitory dysfunction within corticolimbic circuits. Traditional rCBF and positron emission tomography (PET) analyses are regional and not typically analysed within networks of dysfunction. Here, we used a novel individualised-perturbation analysis to investigate whether corticolimbic covariance of rCBF and GABAergic receptor availability are perturbed in individuals at clinical high-risk for psychosis (CHR-P) and first-episode psychosis (FEP), and how the two may be related.

Using simultaneous PET-MRI, we acquired arterial spin labelling (measuring rCBF) and [^11^C]Ro15-4513 PET (measuring α5-GABA_A_R availability) from 24 individuals at CHR-P, 24 healthy controls (HC), and 10 individuals with FEP. Individual covariance deviations were computed for each measure relative to a HC-derived normative reference, with permutation tests assessing group differences. Partial correlations examined hippocampal rCBF-α5-GABA_A_R associations within the network by group and Fisher Z-tests comparing groups.

The magnitude of rCBF covariance deviations were significantly greater in CHR-P and FEP relative to HC for (network: p=.039; hippocampal edges: p=.008) and were predominantly negative (reduced covariance) (network: p=.003; hippocampal edges: p<.001). α5-GABA_A_R covariance deviations were more subtle, with significant negative deviations emerging only in FEP (p=.026; hippocampal edges: p=.010). The relationship between hippocampal rCBF and α5-GABA_A_R availability across several corticolimbic ROIs was significantly different in CHR-P compared to HC (p_FDR_<.050).

Perturbations in corticolimbic rCBF covariance, most prominently at hippocampal edges, are present in at-risk and early psychosis. An altered relationship between hippocampal rCBF and corticolimbic α5-GABA_A_R availability may underlie hippocampal circuit dysfunction in psychosis vulnerability.

## Introduction

Psychosis spectrum disorders are characterised by widespread disruption of coordinated neural activity, which involves the complex integration of diverse neurochemical signals synchronised within networks and across multiple spatiotemporal scales (1,2). Hippocampal-corticolimbic network dysfunction, involving imbalances in excitatory/inhibitory neurotransmission, has emerged as a promising neuroimaging phenotype and potential biomarker in the at-risk and early stages of psychosis (3–7). Two signals offering complementary windows onto this dysfunction are cerebral blood flow (rCBF), a haemodynamic measure reflecting local metabolic demand (8), and the α5 subunit of the γ-aminobutyric acid (GABA)-A receptor (α5-GABA_A_R), which mediates tonic inhibitory neurotransmission within this circuitry (9). However, these signals have been studied largely in isolation and at the regional rather than network level, limiting sensitivity to detect the distributed network-level alterations expected early in the psychosis neurodevelopmental trajectory (6,7). Here, we address this gap using an individualised structural covariance approach, which quantifies how an individual’s network covariance structure deviates from a normative reference connectome (10). We applied this technique simultaneously to [^11^C]Ro15-4513 positron emission tomography (PET) and arterial spin labelling (ASL) magnetic resonance imaging (MRI) in individuals at clinical high risk for psychosis (CHR-P) and experiencing their first episode of psychosis (FEP).

Individuals at clinical high-risk for psychosis (CHR-P) exhibit hippocampal hyperactivity, indexed by elevated regional cerebral blood flow or volume (rCBF/rCBV) (11–13). These elevations track structural MRI changes in CHR-P individuals who subsequently develop psychosis (14,15), are associated with adverse functional outcomes (16), and normalise in individuals whose presenting CHR-P symptoms later resolve (13). Convergent preclinical and post-mortem evidence implicates GABAergic interneuron dysfunction as a proximal cause (17) For instance, preclinical evidence suggests that experimental loss of these interneurons leads to hippocampal dysrhythmia and hyperactivity, in turn propagating aberrant signals through efferent circuitry (18,19). This dysregulation is hypothesised to drive abnormal dopaminergic activity in the midbrain and striatum, which may lead to positive psychotic symptoms, and altered basolateral amygdala-cortical activity, associated with negative psychotic symptoms (14,18,20). However, *in vivo* magnetic resonance spectroscopy (MRS) measuring bulk GABA have yielded inconsistent results in CHR-P and psychosis spectrum disorders (21), likely reflecting both methodological challenges in resolving the GABA signal at conventional field strengths (1.5 and 3 Tesla) and the inherent complexity of the GABA system itself, which involves multiple cellular and receptor subtypes (22). A receptor-specific approach may therefore be important in discerning the GABAergic contributions to corticolimbic circuit dysregulation.

The α5 subunit of the GABA_A_ receptor (α5-GABA_A_R) is a strong candidate marker for dysregulation in corticolimbic circuitry. α5-GABA_A_Rs are preferentially expressed in the hippocampus (23), where they are found extrasynaptically, mediate tonic inhibition, and play a central role in cognition, learning, and memory (9,24,25). In a neurodevelopmental disruption rodent model relevant to psychosis, positive allosteric modulation at α5-GABA_A_R reduced both hippocampal hyperactivity and dopaminergic signalling (26–29). This evidence suggests that α5-GABA_A_R are a potential target for ameliorating hippocampal hyperactivity in CHR-P and early in psychosis spectrum disorders (30). α5-GABA_A_R can be studied using positron emission tomography (PET) with the radiotracer [^11^C]Ro15-4513, which has high selectivity for the α5-GABA_A_R (31). Prior [^11^C]Ro15-4513 PET studies in the psychosis spectrum have been limited to regional analysis; one found no group differences in the hippocampus or prefrontal cortex between 11 individuals with schizophrenia and 12 healthy control participants (HCs) (32), while another study found lower hippocampal α5-GABA_A_R availability in 10 antipsychotic-free individuals with schizophrenia compared with 10 HCs, but not in medicated patients (33). Whether the hippocampal α5-GABA_A_R signal reflects broader corticolimbic network organisation, and whether such network-level organisation is disrupted before frank psychosis onset, has not yet been determined.

The regional focus of prior work is itself a limitation to address. The pathophysiological significance of hippocampal hyperactivity is in its propagation through corticolimbic circuitry to drive downstream dysfunction (7). Group mean regional comparison cannot capture this distributed organisation, and collapsing each clinical group into a single mean obscures the heterogeneity of the group, a core feature of CHR-P (34). Individualised structural covariance analysis addresses these limitations by yielding a per-participant index of network disorganisation (10,35). By constructing a normative reference connectome from HCs and quantifying each individual’s covariance deviation from it, the approach replaces group-mean comparison with a participant-level deviation score, which can then be examined for distributed shifts at the group-level or related to individual clinical features. Applied to structural MRI, this approach has revealed individualised morphometric deviations in schizophrenia (35) and CHR-P (36). It has also recently been extended to molecular imaging of 6-[^18^F]fluro-L-dopa PET (10), but not to perfusion imaging, and not in a multimodal study examining whether neurochemical and haemodynamic perturbation structure relate within the same network.

The present study represents to our knowledge, the first application of this approach to functional and neurochemical data together to identify early circuit-level perturbations linked to psychosis risk and onset. Using simultaneous [¹¹C]Ro15-4513 PET/MRI, we addressed two primary questions in CHR-P. First, whether corticolimbic covariance structure, network-wide and hippocampus-specific, is disrupted in CHR-P for rCBF and α5-GABA_A_R, and in which direction. Second, whether the two modalities are related, specifically whether rCBF and α5-GABA_A_R covariance deviations are correlated, and whether any coupling between regional rCBF in the hippocampus and α5-GABA_A_R availability across the other regions of the corticolimbic network differ between CHR-P and HC. We hypothesised that (1) CHR-P individuals would show disrupted corticolimbic network covariance for both rCBF and α5-GABA_A_R, reflected in negative covariance deviations relative to HC; and (2a) rCBF and α5-GABA_A_R covariance deviations would be correlated, reflecting a shared mechanism of corticolimbic circuit disorganisation, and (2b) the relationship between hippocampal rCBF and α5-GABA_A_R would be altered in the clinical group. As exploratory analyses, we examined whether covariance deviations were associated with clinical symptom severity, and we repeated all primary analyses in a smaller FEP sample (n=10) to test whether the CHR-P pattern extended to early psychosis.

## Methods

### Participants

Twenty-four CHR-P (mean ± SD = 24.97 ±4.35 years, 15 female), 10 FEP (mean ±SD = 29.05 ±3.64 years, seven female), and 24 HC participants (mean ± SD = 25.2 ±4.4, 15 female) were recruited through Outreach and Support in South-London (OASIS) and early intervention services within the South London and Maudsley (SLaM) National Health Service (NHS) Foundation Trust (CHR-P and FEP groups) and via public advertisement (HC group). Full sample characteristics and recruitment criteria have been reported in detail elsewhere (37). Briefly, CHR-P participants met Comprehensive Assessment of At-Risk Mental States (CAARMS) criteria (38) and were antipsychotic naïve. FEP participants met an ICD-10 diagnosis of psychosis (F20-F29 or 31) within two years of their scanning visit and at least moderate severity on a positive symptom dimension of the Positive and Negative Syndrome Scale (PANSS) (39) at assessment. HC participants had no psychiatric history. Ethics approval was obtained through London/Surrey Research Ethics Committee (17/LO/1130). All participants met general eligibility requirements (age 18-40, ≥70 IQ(40), no neurological conditions, no substance dependence, not pregnant or breastfeeding, no MRI contraindications, and no current exposure to GABAergic/glutamatergic compounds) and signed a consent form with capacity to consent.

### Demographic and clinical data

Participants were assessed at King’s College London. Clinical and demographic data were collected, including medical and psychiatric history, substance use, and standardized assessments: Positive and Negative Syndrome Scale (PANSS) for FEP, Comprehensive Assessment of At-Risk Mental States (CAARMS) for CHR-P and HC, Shortened Wechsler Adult Intelligence Scale (WAIS-III)(40) for cognition, Social and Occupational Functioning Assessment Scale (SOFAS) (41) for past-month functioning, Hamilton Anxiety Rating Scale (HAM-A) (42) / Hamilton Depression Rating Scale (HAM-D) (42) for all groups. Blood samples were taken to confirm eligibility for PET scanning (International Normalized Ratio [INR], Activated Partial Thromboplastin Ratio [APTR], Activated Partial Thromboplastin Time [APTT], and prothrombin time). Antipsychotic doses were converted to chlorpromazine equivalents using R 4.2.1 (43). Group comparisons of demographic and clinical data were assessed using chi-square tests (sex) and one-way analysis of variance (ANOVA) (continuous variables). For ethnicity, a permutation-test based on the chi-square statistic (10,000 permutations) was used. All analyses were implemented in Python 3.10.9.

### MRI/PET acquisition

Participants underwent a single-session scan using a General Electric Signa 3T PET/MR scanner with a 12-channel head coil using MP26 software (01 and 02) at the Perceptive (formerly, Invicro) Imaging Centre, London, UK. rCBF data was acquired via 3D pseudo-continuous ASL sequence (multi-shot 3D Fast Spin Echo Stack of Spirals) with a post-labelling delay of 2025ms (TE=11, TR=4854ms, 36 slices, Field of view [FoV]=240mm, slice thickness=3.6mm). CBF maps were calculated in standard physiological units (ml blood/100gm tissue/min) (see supplementary materials for additional details).

The PET acquisition parameters have been reported elsewhere (37). Briefly, α5-GABA_A_R availability was obtained through administration of [^11^C]Ro15-4513 with arterial blood sampling and bolus injection (mean±SD=283.15±73.91 MBq) and 70 minutes dynamic acquisition. For both PET and ASL registration we acquired an T1-weighted Inversion Recovery Fast Spoiled Gradient Recalled (IR-FSPGR) image (1.2×1.05×1.05mm^3^, TE=2.996ms, TR=6.992ms, TI=500ms, flip angle=11.0°, 200 slices, FoV=256mm).

### Image pre-processing

#### Structural T1 imaging

N3 bias correction was applied (44). ROIs were segmented using MAGeT Brain (multiple automatically generated templates of different brains) (45,46). We parcellated subject-space masks of the MNI Allen Human Brain atlas, as well as binarized grey matter masks, which were calculated through Advanced Normalization Tools (ANTs2.5.0 (https://github.com/ANTsX/ANTs). We then extracted eight bilateral ROIs that most closely fitted the hypothesized hippocampus-midbrain-striato-cortical circuit (18)(Figure 1): amygdala, anterior cingulate, associative striatum, hippocampus, midbrain, pallidum, nucleus accumbens (NAc), and orbitofrontal cortex.

**Figure 1.**
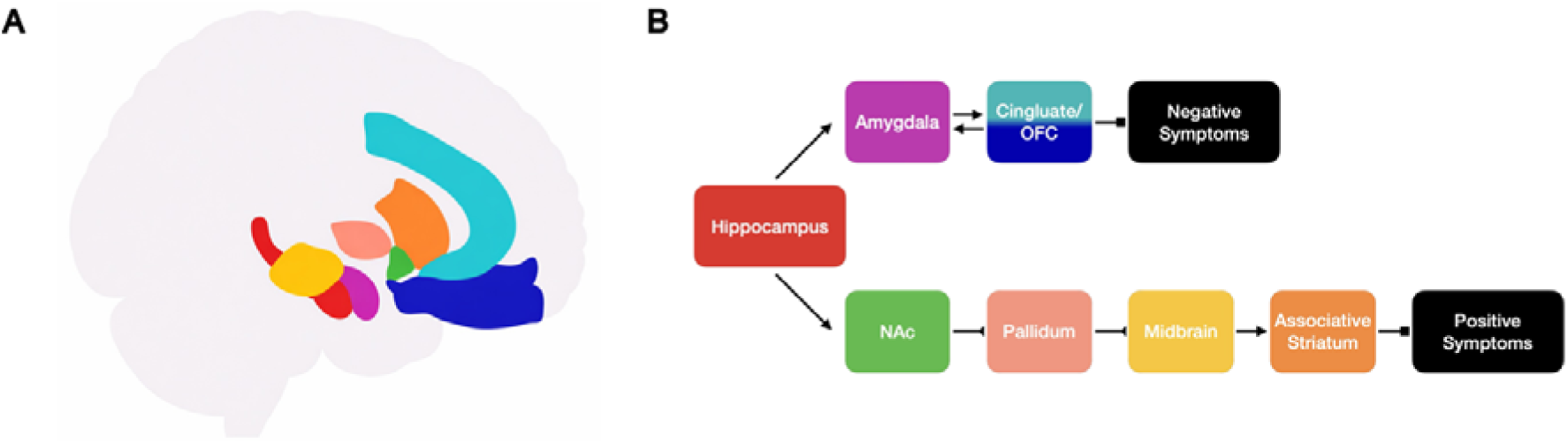
Proposed corticolimbic circuitry underlying positive and negative symptoms, adapted from ref(18). **A** Regions of interest (ROIs) selected for covariance analyses: amygdala (violet), anterior cingulate (cyan), associative striatum (orange), pallidum (pink), hippocampus (red), midbrain/tegmental area (yellow), orbitofrontal cortex (blue). **B** Excitatory and inhibitory circuitry. Forward errors represent excitatory pathways; reverse ended arrows represent inhibitory pathways. Double arrows indicate bidirectional relationship. Square arrows represent a pathway to symptomatology. Nucleus accumbens, NAc. OFC, orbitofrontal cortex. Brain segmentation visualised in surfice 14.6.1 (https://www.nitrc.org/projects/surfice/). Adapted in part from ref(18). Segmentation derived from the Allen Human Reference Atlas(47).

#### ASL preprocessing

Each subject’s quantitative CBF map was sampled in native space. Registered ROIs were resampled to the individual’s CBF map using ANTS and mean rCBF of each ROI was extracted using minc-toolkit-v2/1.9.18 (https://bic-mni.github.io/). One HC was excluded from ASL analysis due to poor data quality. The final sample for ASL was 23 HC, 24 CHR-P, and 10 FEP.

#### PET preprocessing

α*5-GABA_A_R* availability was quantified using volume of distribution (V_T_) by kinetic modelling implemented in MIAKAT 4.3.24 and MATLAB R2017a. Structural images were co-registered to an isotropic, motion-corrected integral image of the PET time series. Continuous and discrete whole-blood data were merged, and plasma time-activity curves (TACs) were generated using a plasma-over-blood model derived from discrete samples. These were then metabolite- and delay-corrected to produce the parent plasma input function, which served as the input for kinetic modelling (see supplementary materials for further details). Four participants were excluded from PET analysis due to quantification errors (one CHR-P, one HC) or issues with data acquisition (one CHR-P, one HC) (see (37) for further details). α*5-GABA_A_R* availability for the midbrain ROI of one CHR participant and one NAc ROI of another CHR-P participant were imputed from the CHR-P group mean due to quantification error. The final sample for the PET and combined analyses was 22 HC, 22 CHR-P, and 10 FEP. To determine the impact of the NAc ROI (larger standard deviations in V_T_ estimations), the main analysis was replicated excluding this ROI (Supplementary materials).

### Statistical analysis

#### Participant demographics and clinical data

Group differences in demographic and clinical variables (age, sex, ethnicity, depression, anxiety, total IQ, SOFAS, and global CBF) were assessed using one-way ANOVA for continuous measures, a chi-square test for sex, and a permutation-based chi-square test (10,000 permutations) for ethnicity. All p-values were corrected for multiple comparisons using the Benjamini-Hochberg false discovery rate (FDR). Significant ANOVAs were followed by post hoc pairwise comparisons using Tukey’s honestly significant difference (HSD) test.

#### Covariance perturbation analyses

##### Individual deviation analyses

For rCBF and α5-GABA_A_R separately, individual Z-score matrices were constructed to quantify deviations in ROI-ROI covariance relative to a normative reference group (HC) (10,35). For each modality, the reference matrix was computed using partial Pearson’s correlations across the eight corticolimbic ROIs (Figure 1) adjusting for age and sex. Additional adjustment for global CBF was performed for rCBF only. For each clinical participant, an individual covariance perturbation matrix was generated by iteratively adding them to the HC reference network. The difference between this augmented connectivity matrix and the original reference matrix was calculated and standardised into a z-score matrix, reflecting the magnitude and direction of covariance change at each ROI pair (edge) relative to the normative reference. Mean absolute deviation (|Z|)-scores were calculated across all edges (network-wide) and for hippocampal edges specifically, indexing overall magnitude of perturbation irrespective of direction. Additionally, for each subject, a mean signed z-score was calculated across all edges (network-wide) and for hippocampal edges specifically, alongside the mean |Z|-score. The signed Z-score captures the net direction of covariance deviation relative to the HC reference, with negative values indicating reduced network covariance and positive values indicating increased covariance. Finally, a medication sensitivity analysis was conducted by rerunning analysis in only unmedicated CHR-P individuals (see Supplementary Materials). Group differences in covariance perturbation scores were assessed using one-tailed permutation tests (10,000 permutations), in which mean differences between groups were compared to a null distribution generated by randomly permutation of group labels, with statistical significance determined as p_FDR_<0.05. Tests were conducted for network-wide and hippocampal-edge specific perturbations separately. CHR-P vs HC was prespecified as primary, FEP vs HC was treated as an exploratory comparison group given the small sample size.

##### Multimodal rCBF and α5-GABA_A_R network covariance correlations

Intraclass correlation coefficients (ICC[2,1]; two-way random effects, absolute agreement) were used to assess the correspondence between network-wide and hippocampal signed perturbation scores in the subsample with complete α5-GABA_A_R and rCBF data (n=22 CHR, 10 FEP). HC were used to construct the reference matrix and therefore were not assessed with ICC. To further examine whether hippocampal rCBF and α5-GABA_A_R availability was associated across the corticolimbic network, partial Pearson’s correlations were computed between hippocampal rCBF and α5-GABA_A_R binding at each network ROI separately within each group, controlling for age, sex, and global CBF. Fisher Z-tests compared correlation strengths between groups (CHR-P vs HC primary; FEP vs HC exploratory), with FDR p<0.05 correction applied across ROIs within each comparison, separately for each modality.

#### Exploratory and sensitivity analyses

##### Symptom correlations

Spearman’s correlations examined associations between |Z| and signed perturbation scores and positive and negative symptom severity (CAARMS), cognitive ability (WAIS-III), global functioning in the past month (SOFAS), anxiety and depression (HAM-A and HAM-D). For FEP, the small sample size (n=10) was insufficient to support reliable symptom correlations and therefore symptom correlations were not performed. Correlations were conducted separately for each modality (p_FDR_<0.05).

##### Regional group differences by modality

Group differences in regional rCBF and α5-GABA_A_R availability were separately assessed using Mann-Whitney U tests with rank-biserial correlation as an effect size measure (p_FDR_<0.05).

All statistical tests were conducted in Python 3.10.9 using SciPy and statsmodels.

## Results

### Participant demographics and clinical data

Demographic/clinical characteristics of the full sample are summarized in Table 1 (Supplementary Table 1 for PET subsample). The groups did not differ in sex distribution or age (both p_FDR_>0.05). However, ethnicity differed significantly across groups (p_FDR_<0.001), driven by a greater proportion of Asian individuals in the HC group relative to the clinical groups. IQ differed across groups (p_FDR_=0.021), with post-hoc Tukey tests indicating significantly lower IQ in FEP compared to HC (mean difference =-20.95, p=0.019), but no significant pairwise differences between CHR-P and HC or CHR-P and FEP. Global functioning also differed across groups (p_FDR_<0.001), with both CHR-P and FEP scoring significantly lower than HC but not differing from each other. Depression and anxiety scores differed significantly across all three groups (both p_FDR_<0.001): both CHR-P and FEP reported higher symptoms than HC (all p<0.05), and CHR-P scored higher than FEP on both measures (depression: p=0.047; anxiety: p=0.046). Global CBF did not survive FDR correction (p_FDR_=0.053).

**Table 1.**
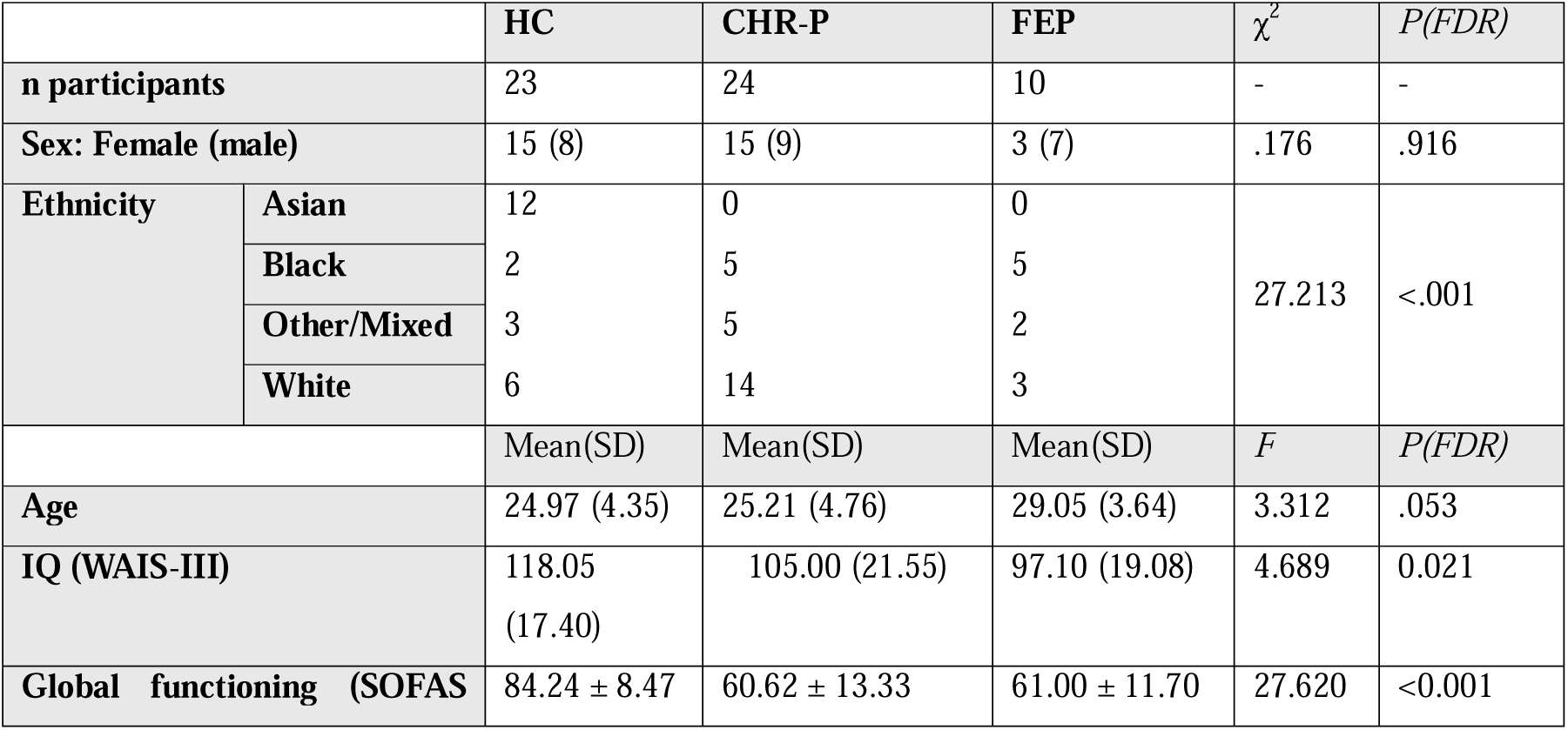
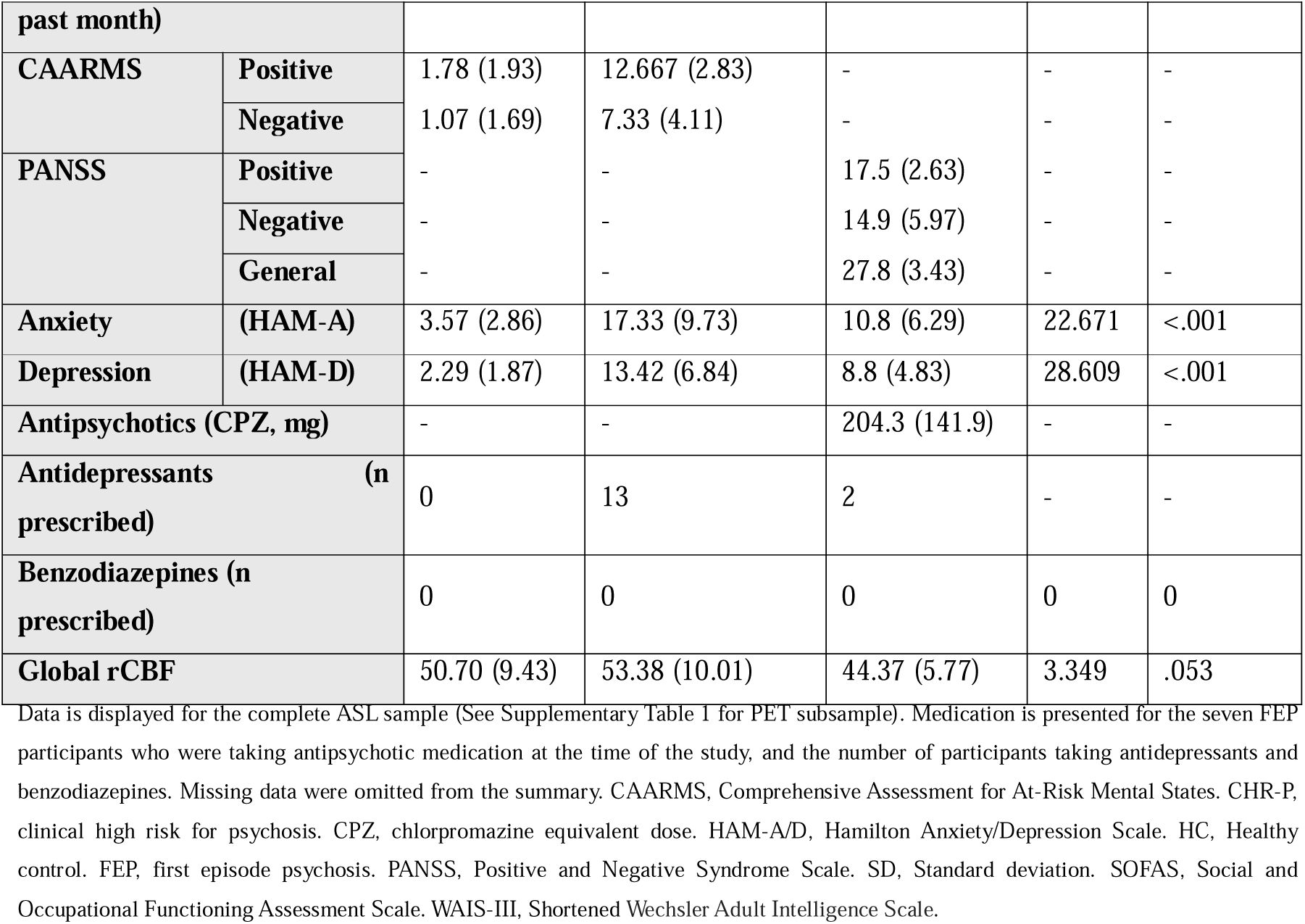
Participant demographic information.

### rCBF covariance perturbations

CHR-P and FEP individuals showed rCBF covariance deviations across multiple corticolimbic network edges, most prominently at hippocampal edges involving the associative striatum (CHR-P and FEP), anterior cingulate (FEP), and amygdala (FEP) (Figure 2A/B). Mean |Z| covariance deviations were significantly greater in CHR-P than HC both network-wide (p=.039, d=0.544) and at hippocampal edges (p=.008, d=0.748; Figure 2C/D, Supplementary Table 2). Signed covariance deviations, which reflect the net direction of covariance change relative to HC, were significantly more negative in CHR-P compared to HC network-wide (p=.003, d=-0.835) and at hippocampal edges (p<.001, d=-1.198). Exploratory analyses in FEP showed a consistent pattern, with significantly greater |Z| deviations network-wide (p=.012, d=0.745) and at hippocampal edges (p<.001, d=1.052), and more negative signed deviations network-wide (p=.026, d=-1.739) and at hippocampal edges (p<.001, d=-1.502), relative to HC. Across both modalities and all perturbation metrics, FEP showed consistently larger effect sizes than CHR-P. NAc sensitivity analysis closely resembled the main results (Supplementary Figure 2). To test whether these effects may be due to medication, we reran the perturbation analysis in only CHR-P who were unmedicated (n=9). Although a small group, |Z| deviations were significantly greater than HC for both across the network and in hippocampal edge, though negative signed Z deviations were no longer significant (Supplementary Figure 4).

**Figure 2.**
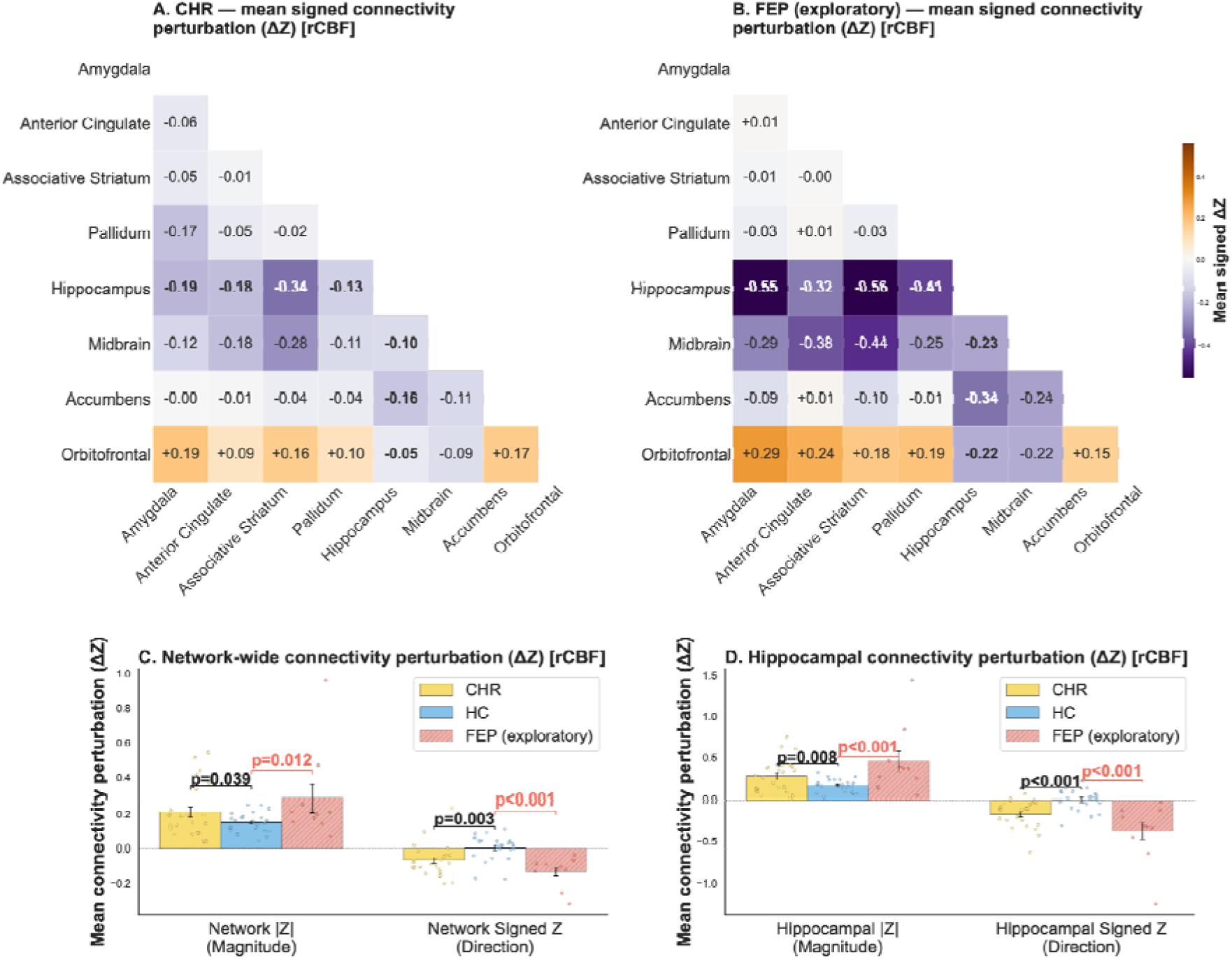
rCBF covariance perturbation analyses. A) Mean signed rCBF covariance deviation matrix for CHR-P relative to the HC reference network; values reflect mean edge-level perturbation z-scores, with positive values indicating increased covariance and negative values indicating decreased covariance relative to HC. Hippocampal edges are shown in bold. B) As in A, for FEP (exploratory). C) Mean |Z| and signed covariance deviation z-scores across the entire network and for hippocampal edges specifically, for CHR-P (primary) and FEP (exploratory). Brackets indicate pairwise permutation test results; exploratory FEP brackets shown in red. D) As in C, for hippocampal edges only. Significance of pairwise permutation tests indicated as ** p<.01, * p<.05, FDR-corrected. CHR-P, clinical high-risk for psychosis; HC, healthy control; FEP, first-episode psychosis.

### α5-GABA_A_R covariance perturbations

α5-GABA_A_R covariance deviations were observed across multiple network edges in both CHR-P and FEP, most prominently between the hippocampus and midbrain in CHR-P, and between the hippocampus and amygdala, hippocampus and associative striatum, orbitofrontal cortex and amygdala, orbitofrontal cortex and associative striatum, and amygdala and midbrain in FEP (Figure 3A/B). Mean |Z| covariance deviations did not differ significantly from HC across the whole network or at hippocampal edges in either group (all p>.10; Figure 3C/D, Supplementary Table 3). Signed covariance deviations were significantly more negative in FEP relative to HC both network-wide (p=.026, d=-0.849) and at hippocampal edges (p=.010, d=-0.958). In CHR-P, signed deviations showed a consistent but non-significant pattern network-wide (p=.157, d=-0.318) and at were at trend-level in hippocampal edges (p=.067, d=-0.465) with medium effect sizes, suggesting the analysis may be underpowered to detect differences of this magnitude. NAc sensitivity analysis closely resembled the main results (Supplementary Figure 3). The α5-GABA_A_R perturbation relationships remained nonsignificant in unmedicated CHR-P (Supplementary Figure 4).

**Figure 3.**
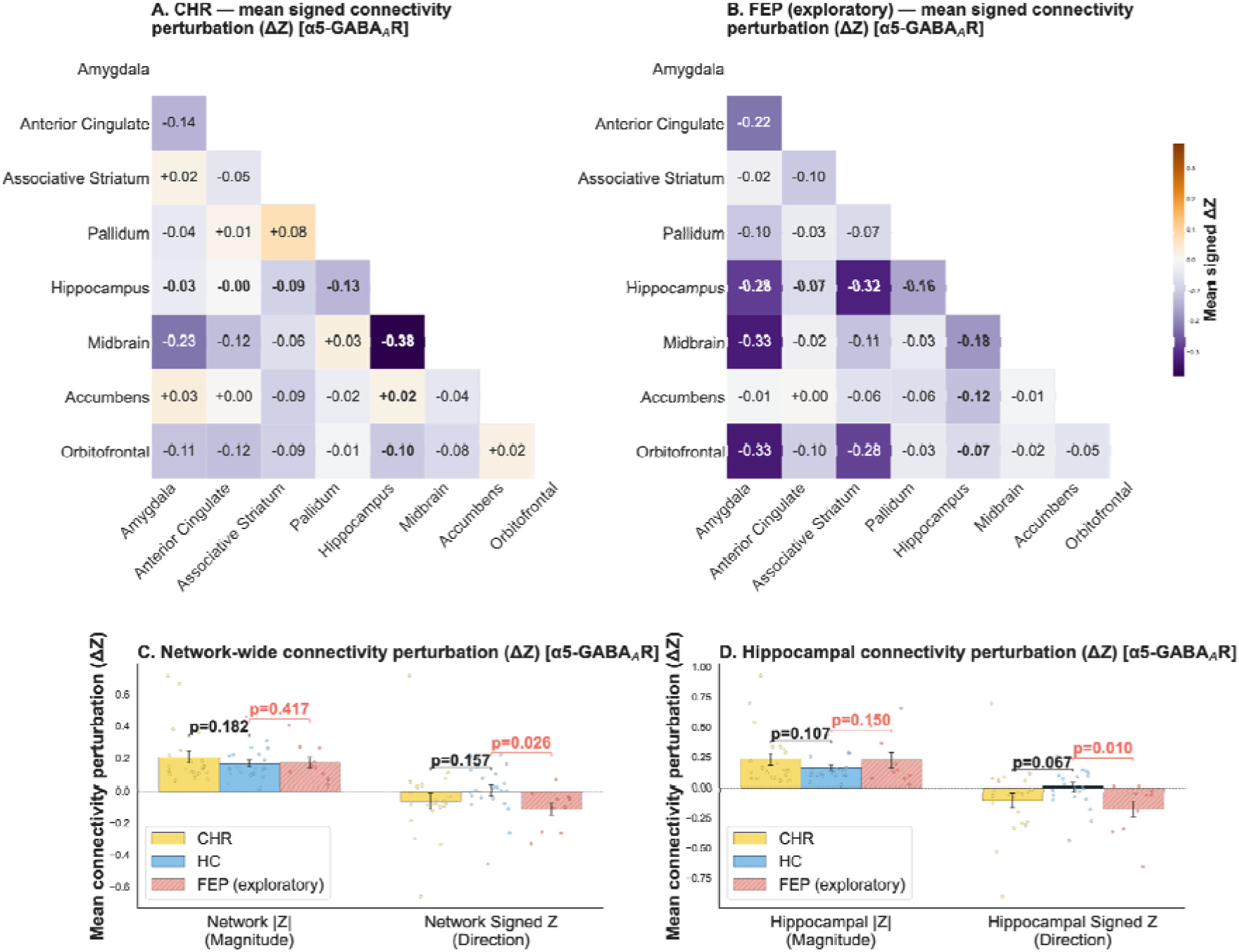
α5-GABA_A_R covariance perturbation analyses. A) Mean signed α5-GABA_A_R covariance deviation matrix for CHR-P relative to the HC reference network; values reflect mean edge-level perturbation z-scores, with positive values indicating increased covariance and negative values indicating decreased covariance relative to HC. Hippocampal edges are shown in bold. B) As in A, for FEP (exploratory). C) Mean |Z| and signed covariance deviation z-scores across the entire network and for hippocampal edges specifically, for CHR-P (primary) and FEP (exploratory). Brackets indicate pairwise permutation test results; exploratory FEP brackets shown in red. D) As in C, for hippocampal edges only. Significance of pairwise permutation tests indicated as ** p<.01, * p<.05, FDR-corrected. CHR-P, clinical high-risk for psychosis; HC, healthy control; FEP, first-episode psychosis.

### Relationships between rCBF and α5-GABA_A_R

#### Covariance perturbation patterns

ICC between rCBF and α5-GABA_A_R network-wide and hippocampal signed perturbation scores in CHR-P and exploratory in FEP were not significant for either signed- or |Z|-score values (CHR-P: network signed Z ICC=-0.092 [95% CI: -0.520–0.350], p=.654; hippocampal signed Z ICC=-0.046 [95% CI: -0.470–0.380], p=.580; FEP: network signed Z ICC=0.024 [95% CI: -0.680–0.640], p=.474; hippocampal signed Z ICC=-0.095 [95% CI: -0.570–0.510], p=.623; see supplementary materials for nonsignificant |Z| tests).

#### Hippocampal rCBF and regional α5-GABA_A_R availability

To examine whether the relationship between hippocampal rCBF and regional GABA_A_R availability, we tested partial correlations between hippocampal rCBF and α5-GABA_A_R availability across the seven corticolimbic ROIs (Figure 4A). In HC, hippocampal rCBF negatively correlated with α5-GABA_A_R in six of seven regions (r range =-.618-.693; p_FDR_<0.05), with no significant association in the pallidum (r=-.418, p_FDR_=0.748). These correlations were absent in CHR-P (r range =-.062-.254; all p_FDR_>0.05). Fisher’s Z tests confirmed significant group differences between HC and CHR-P in the same six regions (amygdala: Z=+2.34, p_FDR_ =.027; anterior cingulate: Z=+2.86, p_FDR_=.010; associative striatum: Z=+2.03, p_FDR_ =.049; hippocampus: Z=+2.86, p_FDR_=.010; midbrain: Z=+3.17, p_FDR_ =.010; orbitofrontal: Z=+2.62, p_FDR_ =.015, pallidum: Z=+1.29, p_FDR_ =.197; Figure 4B). In exploratory analysis of the FEP group, hippocampal rCBF–α5-GABA_A_R correlations were similarly absent across all regions (r range: -0.365 to -0.035, all p_FDR_>.315), though not significantly different from HC (all p_FDR_>.315).

**Figure 4.**
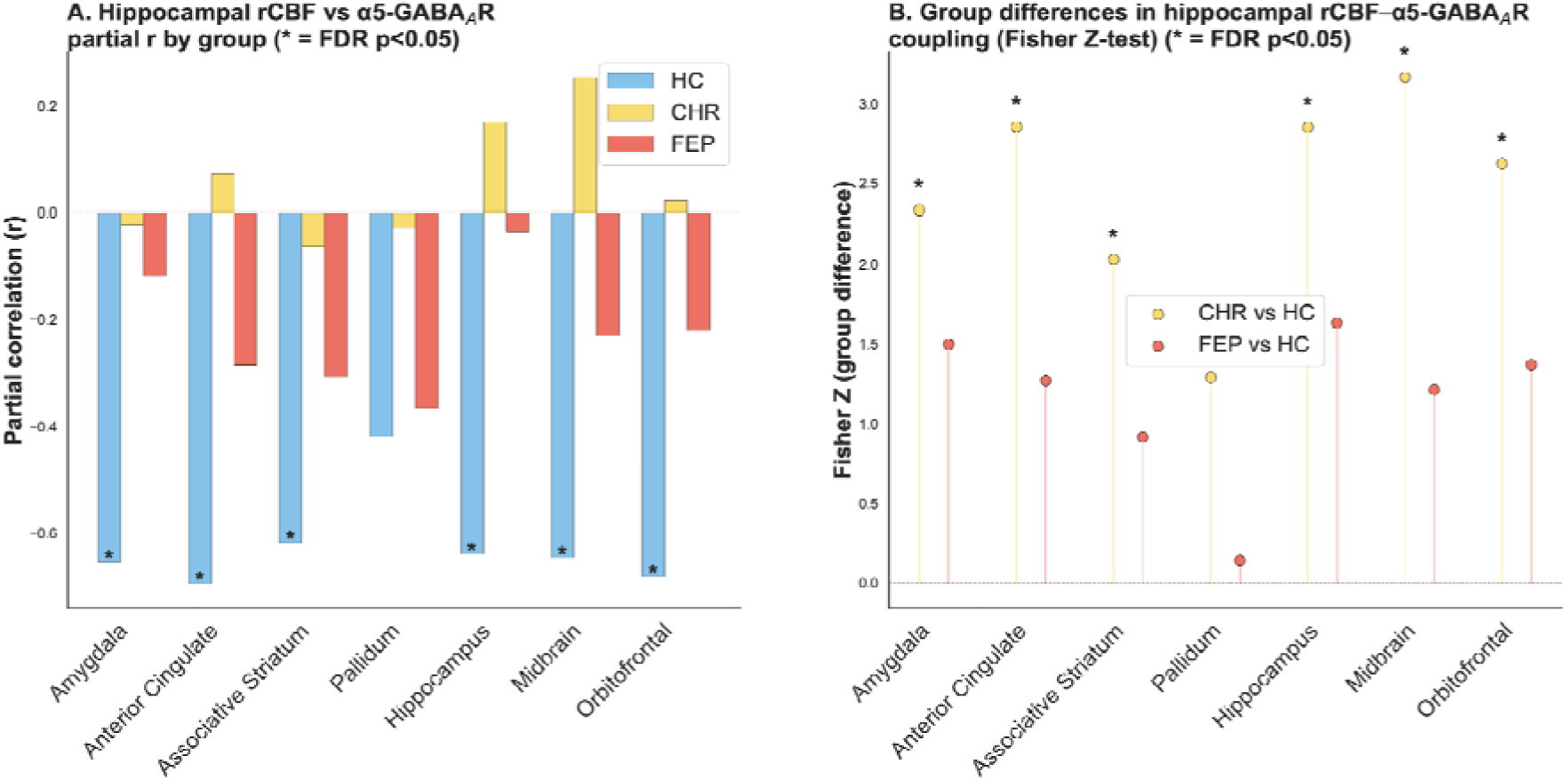
Hippocampal rCBF relationship with α5-GABA_A_R availability across the corticolimbic network. Partial correlations between hippocampal rCBF and α5-GABA_A_R availability at each corticolimbic ROI, computed separately within each group controlling for age, sex, and global CBF. A) Partial correlation coefficients (r) for HC, CHR-P, and FEP. The scatterplots for these partial correlations are shown in Supplementary Figure 5. B) Fisher Z-statistics quantifying the difference in hippocampal rCBF-α5-GABA_A_R correlation strength between clinical groups and HC. Asterisks denote FDR-corrected significance (*p_FDR_*<.05). CHR-P, clinical high risk for psychosis; HC, healthy control; FEP, first episode of psychosis.

### Exploratory and sensitivity analyses

#### Symptom correlations

Spearman correlations between rCBF and α5-GABA_A_R covariance perturbation scores and clinical measures are shown in Supplementary Figure 1. No significant associations were found between signed covariance deviations and CAARMS positive or negative symptoms for either rCBF (all *p_FDR_*>.911) or α5-GABA_A_R (all *p_FDR_*>.213). A nominally significant negative association between rCBF network |Z| and CAARMS positive symptoms (r=−0.523, p=0.009 uncorrected) did not survive FDR correction (*p_FDR_*=.106). No further significant rCBF associations were observed for any clinical measure (all *p_FDR_*>.911). For α5-GABA_A_R perturbation scores, CAARMS symptom associations were non-significant (all *p_FDR_*>.224). However, greater α5-GABA_A_R absolute deviation magnitude (network |Z|) was significantly associated with lower cognition (r=−0.576, *p_FDR_*=.039), higher anxiety (r=+0.551, *p_FDR_*=.039) and depression symptoms (r=+0.539, *p_FDR_*=.039), indicating that greater overall network disorganisation irrespective of direction was associated with lower cognitive and higher affective symptom scores. Depression symptoms additionally showed a significant negative association with α5-GABA_A_R signed Z (r=−0.520, *p_FDR_*=.039), indicating that more negative covariance deviations were specifically associated with higher depression symptoms. A trend-level negative association was observed between α5-GABA_A_R network |Z| and current global functioning (SOFAS: r=−0.466, *p_FDR_*=.070).

#### Regional group differences in rCBF and α5-GABA_A_R

There were no significant between-group differences in either rCBF or α5-GABA_A_R availability in the across any ROIs (Supplementary Materials), but there were trend-level rCBF elevations in FEP.

## Discussion

The main finding of this study was that rCBF covariance within a corticolimbic network is perturbed in individuals at CHR-P and those with FEP relative to a normative HC reference group, with effects driven by negative deviations reflecting reduced network covariance. As hypothesized, covariance perturbations were prominent at hippocampal edges and significantly different from the reference group, with effect sizes for hippocampal edges consistently larger than for network-wide measures across both modalities, supporting the hippocampus as an important locus in corticolimbic disorganisation. The hypothesis of negative α5-GABA_A_R covariance deviations was supported in the exploratory FEP sample, where deviations were significant both network-wide and at hippocampal edges, and partially supported in CHR-P which showed a directionally consistent but nonsignificant pattern. The hypothesis that rCBF and α5-GABA_A_R covariance perturbation scores would be correlated was not supported, which may indicate that the two signals capture independent aspects of corticolimbic disorganisation. However, the relationship between rCBF in the hippocampus and α5-GABA_A_R availability across corticolimbic ROIs was significantly different in people at CHR-P and HC, with strong negative correlations observed in HC but not CHR-P individuals. Across both modalities and all perturbation metrics, the exploratory FEP sample showed larger effect sizes than CHR-P, which may be indicative of stage-related network disorganisation. Critically, these network-level perturbations occurred in the absence of significant group differences in mean rCBF or α5-GABA_A_R availability, indicating that the individualised covariance perturbation approach captured multivariate group effects that conventional regional analysis did not detect. Overall, these findings provide new empirical support for hippocampal network dysfunction in psychosis vulnerability both in terms of neurophysiology and neurochemistry (3,11), and support the use of the individualized covariance perturbation approach to identify them.

The finding that perturbations in rCBF covariance were most pronounced in hippocampal edges and reflected lower covariance aligns with prevalent theories of psychosis as a dysconnectivity syndrome (48,49), and supports a key role for hippocampal dysfunction in vulnerability for psychosis (3,11,14,17). Subtle disturbances in hippocampal circuitry, potentially induced by stress during development(50), have been proposed to impair network-level coordination required for processes such as neural replay(51), cognitive mapping (1), and ultimately contributing to downstream aberrant striatal dopamine and basolateral amygdala-limbic dysfunction (18). Regarding α5-GABA_A_R covariance perturbations, we observed that these were also predominantly negative deviations reflecting reduced covariance, were significant in FEP, and at trend-level for hippocampal edges in CHR-P individuals. This stage-related effect was evident in the direction rather than the absolute magnitude of covariance deviations, suggesting reduced corticolimbic α5-GABA_A_R coordination with transition to FEP. The direction and magnitude of these effects fit with convergent post-mortem and preclinical evidence for GABAergic interneuron dysfunction in psychosis (7,17,52). Post-mortem studies have reported reductions in the GABA-synthesising enzyme GAD67 in dorsolateral-prefrontal cortex and a functional loss of parvalbumin-positive interneurons in the hippocampus and anterior cingulate (53,54), while preclinical literature centres on loss of hippocampal inhibitory tone, which in turn drives disruption of corticolimbic network coordination and downstream mesolimbic dopaminergic regulation (7). Direct *in vivo* support for GABAergic contribution to hippocampal hyperactivity in CHR-P comes from the recent demonstration that acute diazepam, a nonselective GABA_A_ agonist, normalises hippocampal rCBF in CHR-P (55). However prior benzodiazepine-site PET work using [^11^C]flumazenil reported no consistent binding differences in hippocampus or other regions of interest in schizophrenia (56), although one study in CHR-P found reduced binding potential in the right caudate (57). Likewise prior [^11^C]Ro15-4513 PET research in psychosis reported mixed findings (32,33), and our recent study in CHR-P found no significant regional differences in hippocampal α5-GABA_A_R availability (58). It is possible these findings may highlight effects of antipsychotic medications on regional α5-GABA_A_R density (59,60). *In vivo* MRS studies of bulk GABA in the psychosis spectrum have been inconsistent, with reductions most pronounced in unmedicated patients and in FEP (21), a pattern broadly consistent with the stage-related α5-GABA_A_R effect observed here, though the two measures index different aspects of the GABA system and direct comparison should be made cautiously. The convergence of negative findings across GABAergic PET, MRS, and our own prior regional analysis underscores that GABAergic abnormalities in psychosis spectrum disorders and in CHR-P may be more evident when methods are sensitive to distributed network organisation rather than regional means. Future prospective and longitudinal studies are required to assess how reduced functional covariance of the hippocampus relates to adverse clinical and functional outcomes in these at-risk individuals.

Examining whether these covariance patterns are related, we found that the relationship between regional hippocampal rCBF and corticolimbic α5-GABA_A_R availability was significantly different in people at CHR-P and controls. In HC, there were strong negative correlations between regional hippocampal rCBF and α5-GABA_A_R availability across several corticolimbic ROIs that were not seen in CHR-P individuals. In healthy individuals, it is possible that this negative relationship reflects the normative relationship between rCBF and α5-GABA_A_R but requires replication in larger cohorts. The lack of relationship between hippocampal rCBF and α5-GABA_A_R across the network in CHR-P and FEP may be reflective of excitatory shift in excitatory/inhibitory balance towards excitation. This would be consistent with our recent data-driven neuroreceptomic mapping findings that rCBF alterations in individuals at CHR-P, particularly those who subsequently transitioned to psychosis, significantly tracked the distribution of N-methyl-D-aspartate receptors (61). Further multi-modal research integrating rCBF and PET imaging of excitatory receptors such as NMDA in the same cohort of CHR-P and FEP individuals could help clarify whether a shift in E/I dynamics is responsible for the change in hippocampal rCBF and network α5-GABA_A_R availability in early psychosis.

Exploratory analysis of clinical correlates yielded a mixed picture. rCBF covariance perturbation scores showed no significant associations with clinical measures after FDR correction. α5-GABA_A_R perturbation scores showed no association with positive symptom severity, but greater absolute α5-GABA_A_R deviation magnitude was associated with lower cognitive ability and higher anxiety and depression scores in CHR-P. The association with depression was also evident for signed deviations, where more negative α5-GABA_A_R covariance specifically was related to higher depression symptoms. This directional specificity suggests that the depression association is driven by reduced corticolimbic α5-GABA_A_R coordination, rather than network disorganisation more generally. One tentative interpretation is that rCBF covariance disruption may reflect a relatively uniform characterization of the CHR-P group, while α5-GABA_A_R network organization may more closely track a nonspecific affective/cognitive clinical burden relevant to specific CHR-P profiles. As the CHR-P construct is known to be clinically heterogeneous (34), future work in larger samples may apply data-driven clustering approaches to identify specific CHR-P subgroup(s) in which rCBF and/or GABAergic network organization may be related to clinical burden and outcomes.

To our knowledge, this was the first study to apply covariance perturbation statistics to rCBF and [^11^C]Ro15-4513 PET data from the same individuals. As rCBF data are usually averaged across the ASL acquisition, analyses typically focus on univariate group comparisons of means in separate regions. Here, by applying a novel approach to integrate rCBF data in individualized covariance perturbation analyses relative to a healthy reference group, we were able to capture subtle, multivariate hippocampal network perturbations in CHR-P and FEP at the individual level, which were not detectable via traditional regional case-control comparisons. Indeed, hippocampal rCBF did not differ significantly between groups, consistent with mixed evidence from prior studies (12,13,16,62–64), further underscoring the added sensitivity of the network-level approach. Additionally, we worked in native space, ensuring higher anatomical precision at the individual level (18). Another strength of our study is that all CHR-P participants were antipsychotic naïve, and the sensitivity analysis demonstrated that the rCBF perturbation differences in this group remained significant in unmedicated CHR-P. Finally, while ROIs were selected based on their proposed role in psychosis pathophysiology, these techniques could be further refined to explore any network of interest, using larger normative models, and incorporating other PET tracers potentially related to the rCBF signal.

Our study had several limitations. Although adequately powered for detecting α5-GABA_A_R availability differences (37), the sample size was smaller than in some previous rCBF studies in CHR-P (12,13), which could explain the non-significant regional group differences in hippocampal rCBF. Given the limited sample size of the FEP group, covariance perturbation results should be considered exploratory and require replication in larger cohorts. Additionally, this PET/MRI study was not designed to assess transition to psychosis as an outcome, which could bridge the gap between our understanding of hippocampal circuitry in CHR-P and FEP. A further limitation inherent in using these modalities is the relatively low spatial resolution of ASL and PET, and modelling difficulties for [^11^C]Ro15-4513 PET data led to the exclusion of four participants. Spatial resolution limitations of PET imaging also prevented us from distinguishing between specific hippocampal subregions, such as the anterior subiculum or CA1, hypothesized to drive network dysfunction (7,11,14,17,18). Finally, our multimodal analysis directly comparing edge-to-edge covariance perturbations between rCBF-α5-GABA_A_R may oversimplify underlying biological complexity. While we examined hippocampal rCBF-α5-GABA_A_R coupling at the regional level, more fine-grained region-specific interactions, such as whether low hippocampal α5-GABA_A_R relates to higher rCBF in efferent regions, may be obscured in our analysis and warrants further investigation.

In summary, we identified reduced corticolimbic network covariance in resting perfusion (rCBF) and GABAergic receptor organization (α5-GABA_A_R) in CHR-P and FEP. Furthermore, our findings indicate that individuals at CHR-P show an altered relationship between hippocampal rCBF and α5-GABA_A_R availability across this corticolimbic network. This is important because it supports the notion that hippocampal circuit dysfunction in psychosis vulnerability is related to GABAergic mechanisms. The individualised covariance perturbation approach offers a sensitive, individual-level marker of corticolimbic network dysfunction that may aid future prediction and intervention strategies.

## Data Availability

All data produced in the present study are available upon reasonable request to the authors

## Data Availability

The Python 3.10.9 code used to run the analysis. ASL, PET, and other data used in this project will be made available online upon publication.

## Supplementary Materials

Supplementary material is available online.

## Acknowledgements

SRK was supported by a grant from Mental Health Research UK and the Schizophrenia Research Fund. This research was funded by the Wellcome Trust & The Royal Society [Sir Henry Dale Fellowship 202397/Z/16/Z to GM]. MV is supported by EU funding within the MUR PNRR “National Center for HPC, BIG DATA AND QUANTUM COMPUTING (Project no. CN00000013 CN1), the Ministry of University and Research within the Complementary National Plan PNC DIGITAL LIFELONG PREVENTION - DARE (Project no PNC0000002_DARE), and by Fondo per il Programma Nazionale di Ricerca e Progetti di Rilevante Interesse Nazionale (PRIN), (Project no 2022RXM3H7). JJS is part-funded by the National Institute for Health and Care Research (NIHR) Maudsley Biomedical Research Centre (BRC). This manuscript also represents independent research partly funded by the NIHR Maudsley BRC at South London and Maudsley NHS Foundation Trust and King’s College London. Furthermore, OOD is supported by a grant from NIHR Maudsley BRC at South London and Maudsley NHS Foundation Trust. The views expressed are those of the authors and not necessarily those of the NIHR or the Department of Health and Social Care.

For the purposes of open access, the author has applied a Creative Commons Attribution (CC BY) license to any Accepted Author Manuscript version arising from this submission.

We thank all the participants for their time and commitment to the study and gratefully acknowledge the staff at Invicro, OASIS, and the early intervention services of SLaM for their assistance with recruitment and data acquisition.

## Author contributions

SRK and GM conceived the study. SRK wrote the manuscript, with valuable input from all authors. JJS performed PET modelling, MS and MV developed methods for covariance perturbation analysis, with further methodological contributions and formal analysis by SRK. SRK interpreted the results and made all figures. SRK, PL, AK, NRL, ZA, ADM, JDa, TP, JDo, PFP contributed to recruitment. GM provided supervision and secured funding. SRK was the project administrator.

