## Supplementary Materials for "Corticolimbic network perturbations in clinical high-risk and first-episode psychosis: an arterial spin labelling and [^11^C]Ro15-4513 positron emission tomography study"

**Running title**: rCBF and α5-GABA_A_ receptor hippocampal circuit covariance in psychosis

**Subject terms:** hippocampus; multimodal neuroimaging; arterial spin labelling; positron emission tomography; mental health; network covariance statistics; biomarkers

**METHODS**

**Cerebral blood flow calculation**

Cerebral blood flow maps were calculated in standard physiological units (ml blood/100gm tissue/min) using the following formula:

$$CBF =\frac{\left( 6000 * w \right)}{T1_{a}}. \left( \frac{P}{R} \right).\frac{2ET1_{a}\left( 1 - e^{\frac{-s}{T1_{a}}} \right)}{k}$$

where P represents the signal difference between control and label in the perfusion-weighted image, R is the signal intensity in the reference image, E is the labelling efficiency, s is the labelling duration, T1a is the longitudinal relaxation time of arterial water, w is the post-labelling delay, and k is a scaling constant. The constants used in the reconstruction were: brain–blood partition coefficient = 0.9 mL/g; labelling efficiency E = 0.80; T1a = 1600 ms; labelling duration s = 1500 ms; post-labelling delay w = 2025 ms. Background suppression was applied using four inversion pulses and is accounted for separately in the GE Volume Viewer reconstruction (vxtl_15.0-2.0).

**PET modelling**

TACs for each ROI were extracted from the dynamic PET acquisition using the subject-specific atlas and then input into a two-tissue compartmental model using the brain tissue and blood as the two compartments. The outcome measure from this model is the total volume of distribution (V_T_)_._ Quantification output (brain segmentation, motion correction, left-right image flip, time-activity curves, and parent plasma fraction plots) and tracer kinetic estimates were used for quality checks for each participant.

**Region of interest (ROI) analysis**

To assess group differences and contextualize the covariance perturbation findings, Mann-Whitney U tests compared regional rCBF and α5-GABA_A_R availability between groups (CHR-P vs HC; FEP vs HC) at each of the eight corticolimbic ROIs separately. Effect sizes were calculated as rank-biserial correlation (r). False discovery rate correction (Benjamini-Hochberg) was applied within each modality and comparison across the eight ROIs.

**Sensitivity analyses**

Two sensitivity analyses were conducted to assess the robustness of the primary covariance perturbation findings. First, the primary analysis included nucleus accumbens (NAc) as a ROI despite challenges for V_T_ estimation due to its small size. To assess whether inclusion of this ROI influenced findings, all primary perturbation analyses were repeated using a seven-ROI network excluding the NAc ROI. Second, a medication sensitivity analysis was conducted to assess whether rCBF and α5-GABA_A_R covariance perturbations in CHR-P were attributable to antidepressant medication use. The primary permutation tests were repeated in unmedicated CHR-P individuals only (rCBF: n=11; α5-GABA_A_R: n=9), compared against the full HC reference group. FEP was excluded from this analysis given that 8 of 10 individuals were medicated.

All statistical analyses were performed using statsmodels Python (3.10.9).

**RESULTS**

**Supplementary Table 1.** Participant demographic information for PET subsample

| **Variable** | **HC (n=22)** | **CHR-P (n=22)** | **FEP (n=10)** | **Statistic** | **p value** |
| --- | --- | --- | --- | --- | --- |
| **n participants** | 22 | 22 | 10 | – | – |
| **Sex: Female (male)** | 14 (8) | 15 (7) | 3 (7) | χ²(2)=0.164 | 0.921 |
| **Ethnicity** |  |  |  | χ²(6)=24.396 | <0.001 |
| ─ Asian | 11 (50%) | 0 (0%) | 0 (0%) | – | – |
| ─ Black | 2 (9%) | 5 (23%) | 5 (50%) | – | – |
| ─ Other | 2 (9%) | 4 (18%) | 2 (20%) | – | – |
| ─ White | 7 (32%) | 13 (59%) | 3 (30%) | – | – |
| **Age (years)** | 24.83 (4.39) | 25.11 (4.64) | 29.05 (3.64) | F = 3.570 | 0.035 |
| **CAARMS – Positive** | 1.82 (1.97) | 12.73 (2.68) | – | – | – |
| **CAARMS – Negative** | 0.50 (1.22) | 7.05 (4.37) | – | – | – |
| **PANSS – Positive** | – | – | 17.50 (2.64) | – | – |
| **PANSS – Negative** | – | – | 14.90 (5.97) | – | – |
| **PANSS – General** | – | – | 27.80 (3.43) | – | – |
| **Anxiety (HAM-A)** | 3.75 (2.81) | 17.59 (10.12) | 10.80 (6.29) | F = 18.525 | <0.001 |
| **Depression (HAM-D)** | 2.40 (1.85) | 12.50 (6.38) | 8.80 (4.83) | F = 23.430 | <0.001 |
| **Antipsychotics (CPZ, mg)** | – | – | 204.3 (141.9) | – | – |
| **Antidepressants (n prescribed)** | 0 | 13 | 2 | – | – |
| **Benzodiazepines (n prescribed)** | 0 | 0 | 0 | – | – |

Continuous variables: Mean (SD). Categorical: n (%) or n. CAARMS reported for HC and CHR-P; PANSS for FEP. Global rCBF excluded (ASL-specific measure). Four participants excluded from PET analysis due to quantification or acquisition errors (one CHR-P, one HC each; see Methods).

**Group differences in rCBF and** **α5-GABA_A_R**

No significant regional differences in α5-GABA_A_R availability were observed between CHR-P and HC at any ROI after FDR correction (all p_FDR_=1.000), nor between FEP and HC (all p_FDR_>.638). Similarly, no significant regional rCBF differences were observed between CHR-P and HC (all p_FDR_>.840). For FEP vs HC, several ROIs showed nominally significant rCBF elevations that did not survive FDR correction, including amygdala (r=+0.443, p=.048, p_FDR_=.077), anterior cingulate (r=+0.496, p=.027, p_FDR_=.059), pallidum (r=+0.513, p=.022, p_FDR_=.059), NAc (r=+0.522, p=.020, p_FDR_=.059), and orbitofrontal cortex (r=+0.487, p=.030, p_FDR_=.059), all reflecting higher rCBF in FEP relative to HC. Hippocampal rCBF did not differ significantly between any group pair (CHR-P vs HC: p_FDR_=.840; FEP vs HC: p_FDR_=.129).

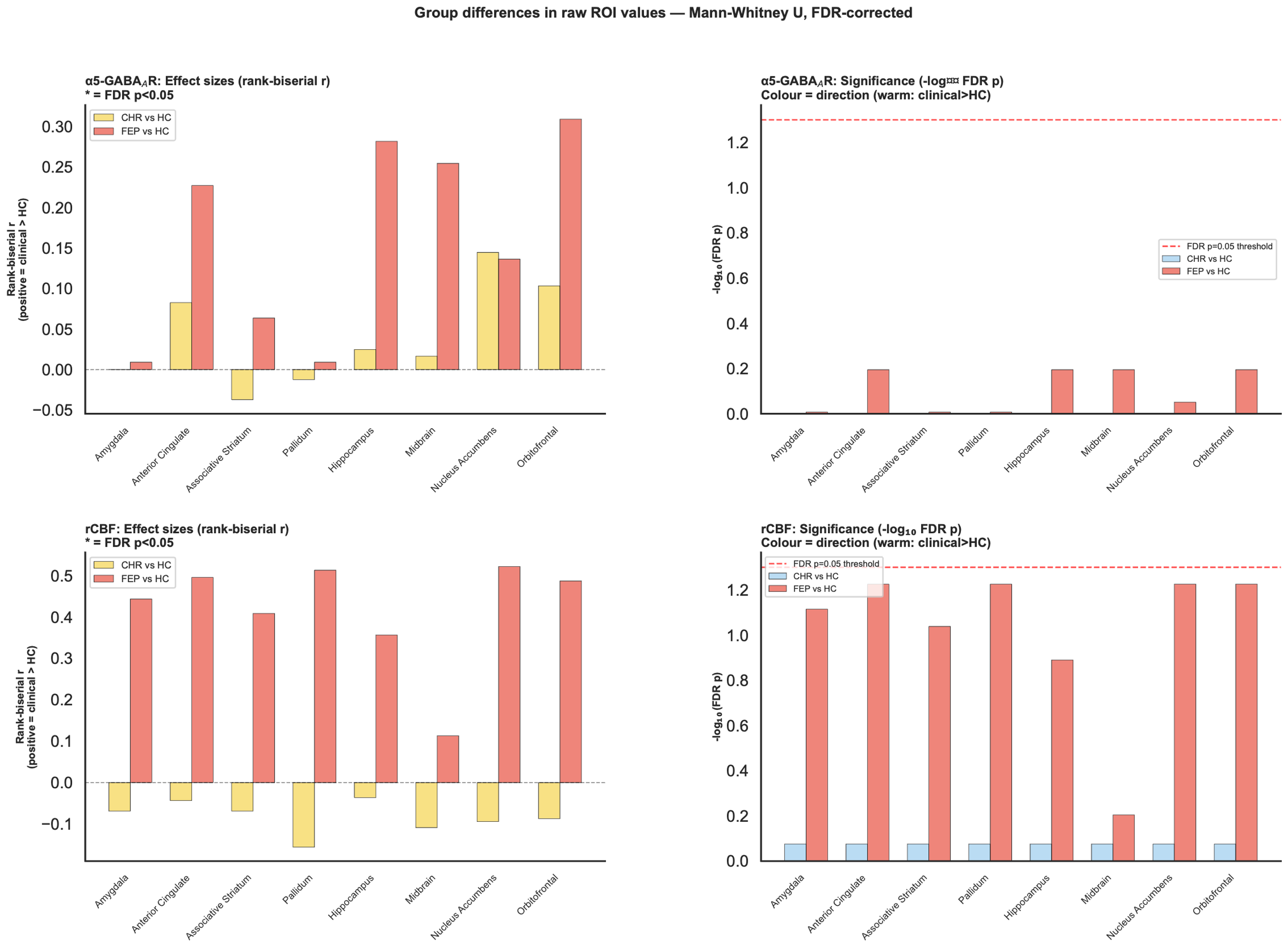

**Supplementary Figure 1. Group differences in mean ROI values.** ASL, arterial spin labelling; CHR-P, clinical high risk for psychosis; HC, healthy control; FEP, first episode of psychosis; rCBF, regional cerebral blood flow.

**Supplementary Table 2.** Primary perturbation analysis results

PRIMARY [CBF]:

| **Metric** | **HC mean** | **HC sd** | **CHR mean** | **CHR sd** | **Difference** | **p value** | **Cohen’s d** |
| --- | --- | --- | --- | --- | --- | --- | --- |
| Network \|Z\| | 0.150933 | 0.055857 | 0.210203 | 0.143554 | 0.059270 | 0.0389 | 0.544152 |
| Network Signed Z | 0.003509 | 0.070338 | -0.063897 | 0.089844 | 0.067407 | 0.0029 | -0.835451 |
| Hippocampal \|Z\| | 0.165189 | 0.072686 | 0.275352 | 0.195219 | 0.110163 | 0.0084 | 0.747887 |
| Hippocampal Signed Z | 0.002388 | 0.116997 | -0.165314 | 0.159773 | 0.167702 | 0.0000 | -1.197633 |
| **Metric** | **HC mean** | **HC sd** | **FEP mean** | **FEP sd** | **Difference** | **p value** | **Cohen’s d** |
| Network \|Z\| | 0.150933 | 0.055857 | 0.284486 | 0.247237 | 0.133553 | 0.0117 | 0.745151 |
| Network Signed Z | 0.003509 | 0.070338 | -0.130906 | 0.083638 | 0.134416 | 0.0000 | -1.739450 |
| Hippocampal \|Z\| | 0.165189 | 0.072686 | 0.457191 | 0.385880 | 0.292002 | 0.0000 | 1.051665 |
| Hippocampal Signed Z | 0.002388 | 0.116997 | -0.375690 | 0.336143 | 0.378078 | 0.0000 | -1.502247 |

PRIMARY [PET]:

| **Metric** | **HC mean** | **HC sd** | **CHR mean** | **CHR sd** | **Difference** | **p value** | **Cohen’s d** |
| --- | --- | --- | --- | --- | --- | --- | --- |
| Network \|Z\| | 0.164876 | 0.095814 | 0.205678 | 0.173886 | 0.040802 | 0.1818 | 0.290641 |
| Network Signed Z | 0.000793 | 0.147885 | -0.061304 | 0.232996 | 0.062097 | 0.1571 | -0.318224 |
| Hippocampal \|Z\| | 0.160783 | 0.114081 | 0.230219 | 0.222067 | 0.069437 | 0.1069 | 0.393334 |
| Hippocampal Signed Z | 0.005658 | 0.158301 | -0.102610 | 0.288681 | 0.108268 | 0.0668 | -0.465062 |
| **Metric** | **HC mean** | **HC sd** | **FEP mean** | **FEP sd** | **Difference** | **p value** | **Cohen’s d** |
| Network \|Z\| | 0.164876 | 0.095814 | 0.172220 | 0.105951 | 0.007344 | 0.4167 | 0.072703 |
| Network Signed Z | 0.000793 | 0.147885 | -0.113597 | 0.120267 | 0.114390 | 0.0262 | -0.848678 |
| Hippocampal \|Z\| | 0.160783 | 0.114081 | 0.223171 | 0.203727 | 0.062389 | 0.1497 | 0.377872 |
| Hippocampal Signed Z | 0.005658 | 0.158301 | -0.171381 | 0.208020 | 0.177039 | 0.0103 | -0.957798 |

**Supplementary Table 3.** Sensitivity analysis without NAc ROI

SENSITIVITY [CBF_sensitivity_no_nacc]:

| **Metric** | **HC mean** | **HC sd** | **CHR mean** | **CHR sd** | **Difference** | **p value** | **Cohen’s d** |
| --- | --- | --- | --- | --- | --- | --- | --- |
| Network \|Z\| | 0.153462 | 0.057448 | 0.229631 | 0.158106 | 0.076169 | 0.0189 | 0.640346 |
| Network Signed Z | 0.002636 | 0.081212 | -0.076182 | 0.094166 | 0.078817 | 0.0018 | -0.896389 |
| Hippocampal \|Z\| | 0.165179 | 0.076669 | 0.286447 | 0.203814 | 0.121268 | 0.0049 | 0.787570 |
| Hippocampal Signed Z | 0.003425 | 0.121668 | -0.166288 | 0.158203 | 0.169712 | 0.0000 | -1.202587 |
| **Metric** | **HC mean** | **HC sd** | **FEP mean** | **FEP sd** | **Difference** | **p value** | **Cohen’s d** |
| Network \|Z\| | 0.153462 | 0.057448 | 0.323925 | 0.273268 | 0.170462 | 0.0023 | 0.863304 |
| Network Signed Z | 0.002636 | 0.081212 | -0.145451 | 0.099636 | 0.148087 | 0.0001 | -1.629268 |
| Hippocampal \|Z\| | 0.165179 | 0.076669 | 0.476087 | 0.379356 | 0.310908 | 0.0000 | 1.136076 |
| Hippocampal Signed Z | 0.003425 | 0.121668 | -0.382116 | 0.324240 | 0.385541 | 0.0000 | -1.574395 |

SENSITIVITY [PET_sensitivity_no_nacc]:

| **Metric** | **HC mean** | **HC sd** | **CHR mean** | **CHR sd** | **Difference** | **p value** | **Cohen’s d** |
| --- | --- | --- | --- | --- | --- | --- | --- |
| Network \|Z\| | 0.153462 | 0.057448 | 0.229631 | 0.158106 | 0.076169 | 0.0189 | 0.640346 |
| Network Signed Z | 0.002636 | 0.081212 | -0.076182 | 0.094166 | 0.078817 | 0.0018 | -0.896389 |
| Hippocampal \|Z\| | 0.165179 | 0.076669 | 0.286447 | 0.203814 | 0.121268 | 0.0049 | 0.787570 |
| Hippocampal Signed Z | 0.003425 | 0.121668 | -0.166288 | 0.158203 | 0.169712 | 0.0000 | -1.202587 |
| **Metric** | **HC mean** | **HC sd** | **FEP mean** | **FEP sd** | **Difference** | **p value** | **Cohen’s d** |
| Network \|Z\| | 0.162674 | 0.093208 | 0.193758 | 0.122347 | 0.031084 | 0.2201 | 0.285808 |
| Network Signed Z | 0.001530 | 0.143623 | -0.137332 | 0.129452 | 0.138862 | 0.0108 | -1.015657 |
| Hippocampal \|Z\| | 0.160045 | 0.110349 | 0.232309 | 0.215114 | 0.072264 | 0.1279 | 0.422710 |
| Hippocampal Signed Z | 0.005815 | 0.154340 | -0.180258 | 0.218414 | 0.186072 | 0.0078 | -0.983936 |

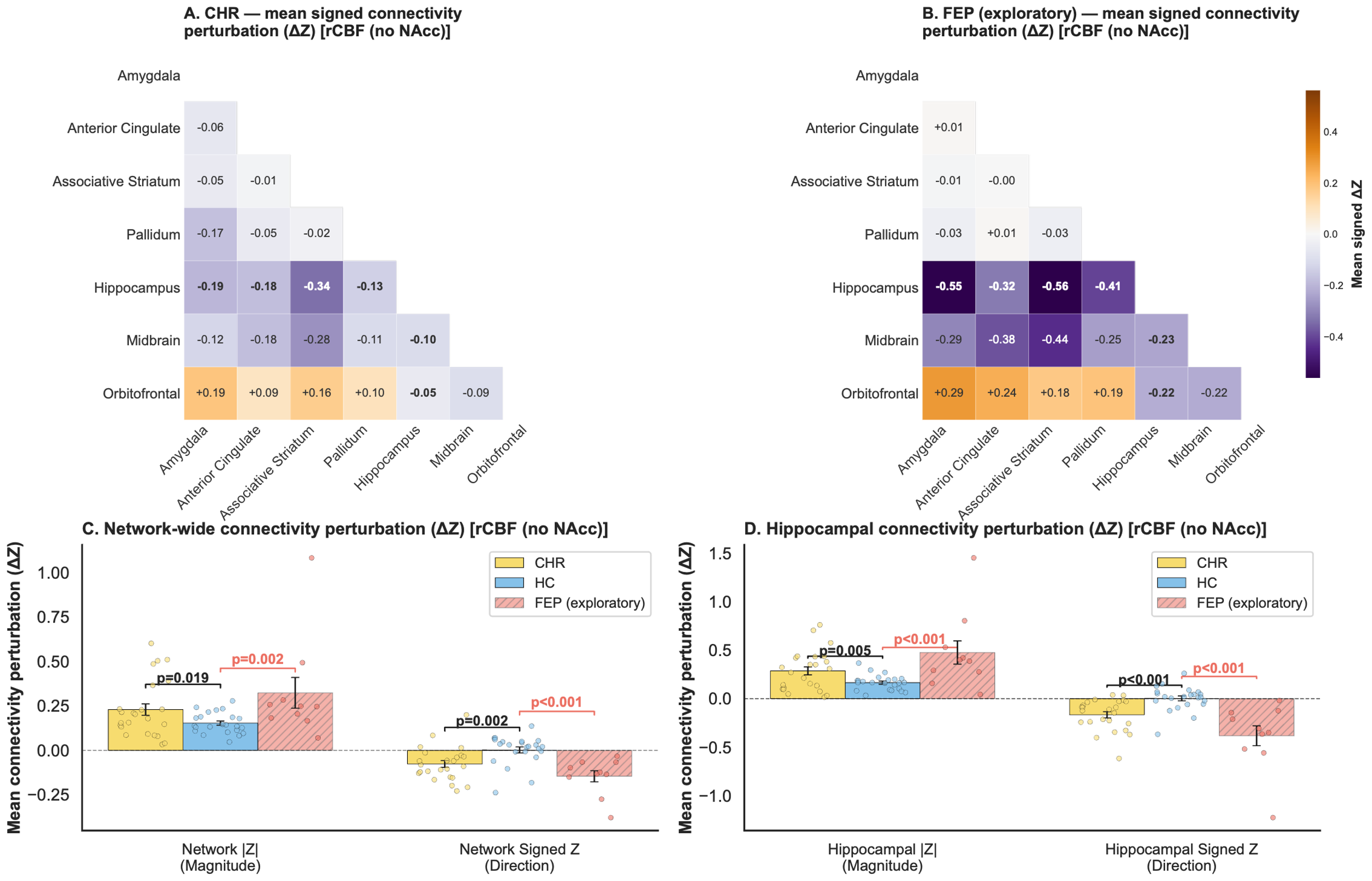

**Supplementary Figure 2. Sensitivity rCBF covariance perturbation analyses (No NAc).** A) Mean signed rCBF covariance deviation matrix for CHR-P relative to the HC reference network; values reflect mean edge-level perturbation z-scores, with positive values indicating increased covariance and negative values indicating decreased covariance relative to HC. Hippocampal edges are shown in bold. B) As in A, for FEP (exploratory). C) Mean signed and |Z| covariance deviation z-scores across the entire network and for hippocampal edges specifically, for CHR-P (primary) and FEP (exploratory). Brackets indicate pairwise permutation test results; exploratory FEP brackets shown in red. D) As in C, for hippocampal edges only. Significance of pairwise permutation tests indicated as ** p<.01, * p<.05, FDR-corrected. CHR-P, clinical high risk for psychosis; HC, healthy control; FEP, first episode of psychosis; NAc, nucleus accumbens; rCBF, regional cerebral blood flow.

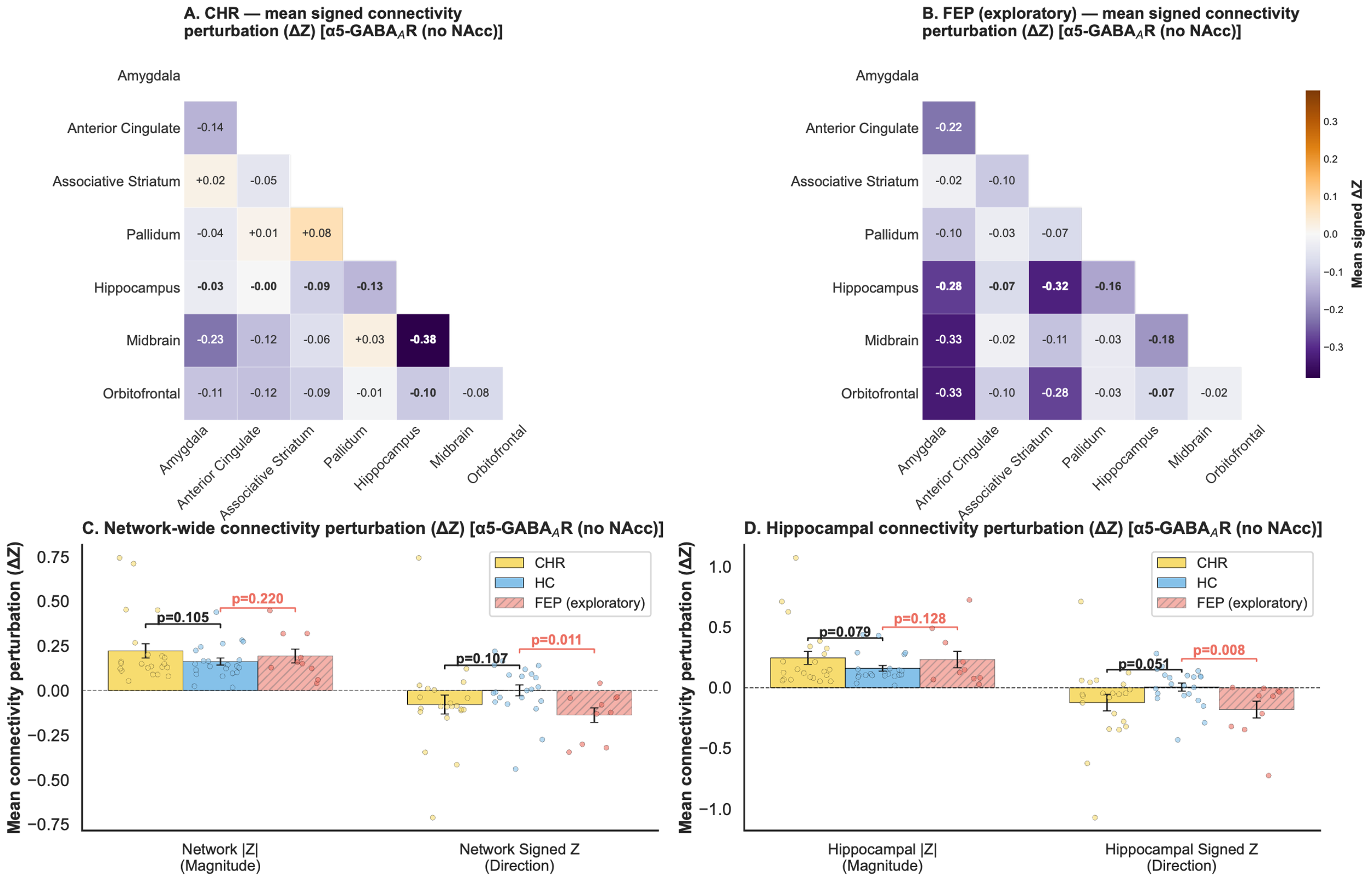

**Supplementary Figure 3.** **Sensitivity** **α5-GABA_A_R covariance perturbation analyses (No NAc).** A) Mean signed α5-GABA_A_R covariance deviation matrix for CHR-P relative to the HC reference network; values reflect mean edge-level perturbation z-scores, with positive values indicating increased covariance and negative values indicating decreased covariance relative to HC. Hippocampal edges are shown in bold. B) As in A, for FEP (exploratory). C) Mean signed and |Z| covariance deviation z-scores across the entire network and for hippocampal edges specifically, for CHR-P (primary) and FEP (exploratory). Brackets indicate pairwise permutation test results; exploratory FEP brackets shown in red. D) As in C, for hippocampal edges only. Significance of pairwise permutation tests indicated as ** p<.01, * p<.05, FDR-corrected. CHR-P, clinical high risk for psychosis; HC, healthy control; FEP, first episode of psychosis; NAc, nucleus accumbens.

**Medication sensitivity analysis**

To test the effect of medication, we isolated the perturbation analysis to only those individuals who were unmedicated. In unmedicated CHR-P, rCBF absolute covariance deviations were significant for both network-wide and hippocampal edges (both p=.038), consistent with the full-sample analysis. Signed Z metrics were non-significant for rCBF (network p=.965, hippocampal p=1.000). For α5-GABAAR, neither signed Z (network p=.951, hippocampal p=.990) nor absolute deviations (network p=.474, hippocampal p=.290) were significant in the unmedicated subsample, though this likely reflects reduced power (n=9 unmedicated CHR-P).

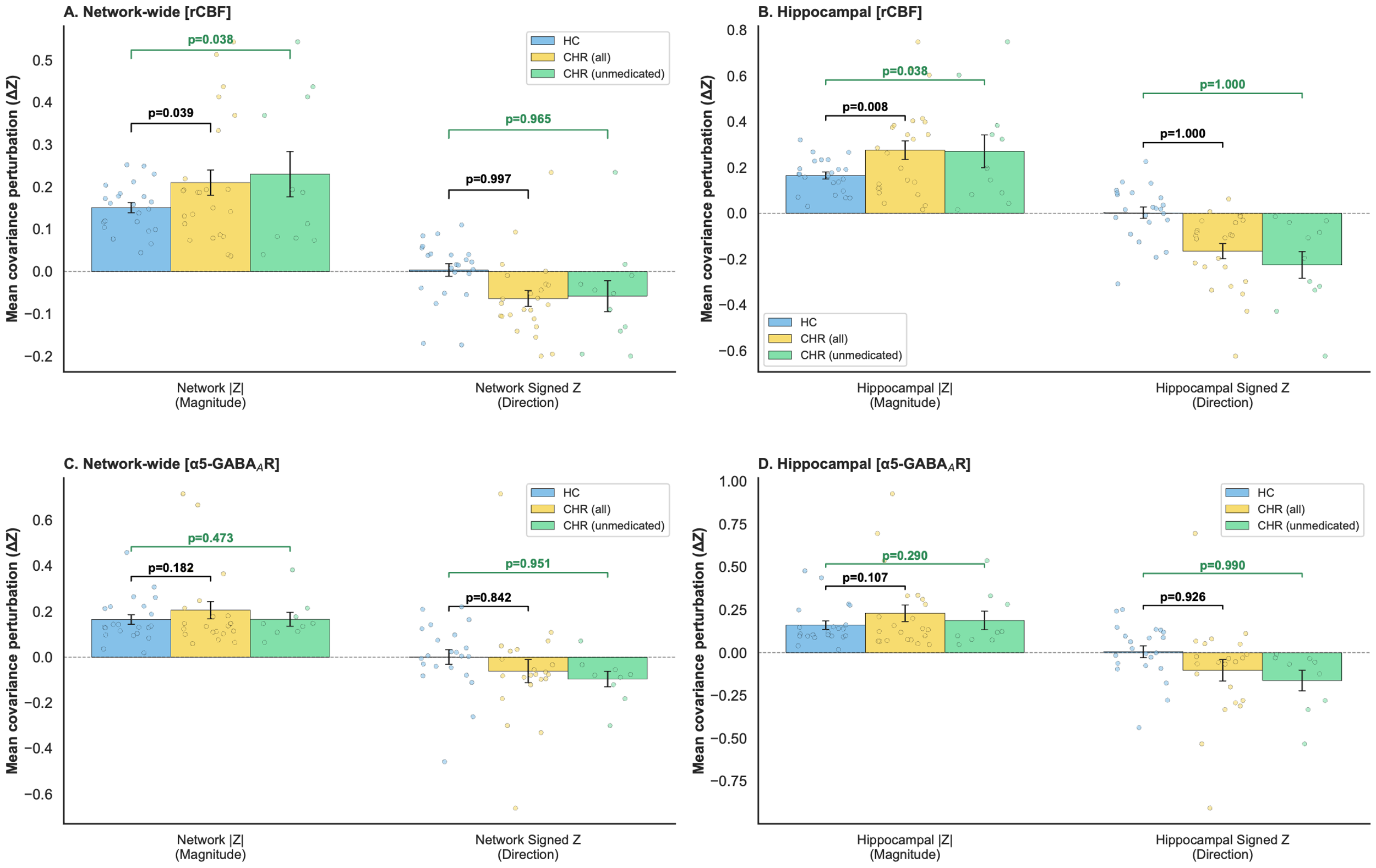

**Supplementary Figure 4. Medication sensitivity analysis for rCBF and α5-GABA_A_R**. CHR-P, clinical high risk for psychosis; HC, healthy control; FEP, first episode of psychosis

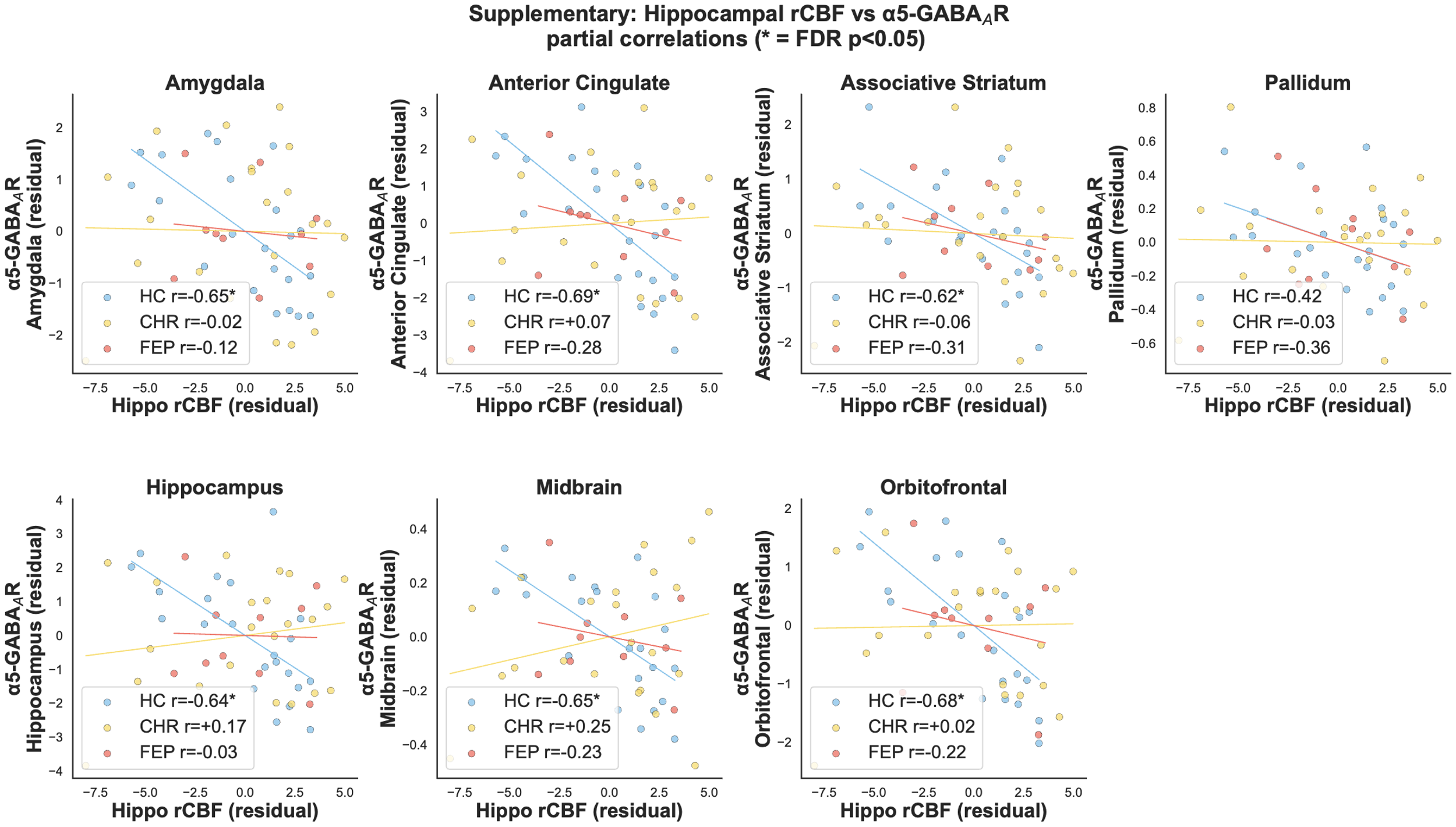

**Supplementary Figure 5. Partial correlation scatterplots of** **hippocampal rCBF relationship with α5-GABA_A_R** **availability** **across the corticolimbic network** CHR-P, clinical high risk for psychosis; HC, healthy control; FEP, first episode of psychosis; Hippo, hippocampus; rCBF, regional cerebral blood flow.
